# Development and Pilot Validation of ABHA-O-SHINE: An AI-Ready Oral Health Risk and Insurance Prediction Framework within the Ayushman Bharat Digital Ecosystem

**DOI:** 10.64898/2026.03.31.26349846

**Authors:** Yash Saxena, Leher Shrivastava

**Author notes:** (Corresponding Author) Dr Yash Saxena, Email id-, Department of Public Health & Community Dentistry, Swargiya Dadasaheb Kalmegh Smruti Dental College & Hospital, Hingna, Nagpur, Maharashtra, India. All Authors declare to have seen and approve the manuscript below.

## Abstract

**Background:** Oral health remains inadequately integrated within the Ayushman Bharat Digital Mission (ABDM), particularly in terms of structured risk assessment and its linkage to insurance-based decision-making. There is a growing need for scalable models that can connect clinical oral health data with digital health systems and support future artificial intelligence (AI)-driven applications.

**Aim:** To develop and pilot test the ABHA-O-SHINE framework for oral health risk prediction and insurance prioritization, with a future scope for AI integration within the Ayushman Bharat Health Account (ABHA) ecosystem.

**Materials and Methods:** A cross-sectional pilot study was conducted among 126 participants attending the outpatient department of Swargiya Dadasaheb Kalmegh Smruti Dental College and Hospital, Nagpur. Participants were selected based on predefined inclusion and exclusion criteria. Data collection included a structured questionnaire and clinical examination using the WHO Oral Health Assessment Form (2013). A composite risk score (0–14) was developed incorporating behavioral and clinical parameters. Participants were categorized into low, moderate, and high-risk groups, and corresponding insurance priority levels were assigned. Statistical analysis included descriptive statistics, Chi-square test, Spearman correlation, and binary logistic regression.

**Results:** The majority of participants were categorized under moderate to high-risk groups. Tobacco use showed a statistically significant association with higher risk levels (p < 0.05). Positive correlations were observed between total risk score and clinical indicators such as DMFT and CPI. Logistic regression analysis identified tobacco use and clinical scores as significant predictors of high-risk categorization.

**Conclusion:** The ABHA-O-SHINE framework demonstrates feasibility in integrating oral health risk assessment with an insurance prioritization model. The framework is designed to be AI-compatible, enabling future automation through machine learning and image-based analysis within the ABDM ecosystem.

## INTRODUCTION

Oral diseases continue to be among the most prevalent non-communicable diseases globally, affecting individuals across all age groups and significantly impacting quality of life. According to the World Health Organization, oral diseases affect nearly 3.5 billion people worldwide, with dental caries and periodontal diseases being the most common conditions [1]. Despite this burden, oral health often remains inadequately integrated into general health systems, particularly in low- and middle-income countries.

In India, the prevalence of dental caries and periodontal disease remains high, influenced by behavioral, socioeconomic, and environmental factors [2]. Existing healthcare delivery systems largely focus on treatment rather than preventive and risk-based approaches, leading to increased long-term healthcare costs and unmet needs.

The Ayushman Bharat Digital Mission (ABDM) represents a major step toward digitization and integration of healthcare in India through the Ayushman Bharat Health Account (ABHA), enabling longitudinal health records and improved healthcare access [3,4]. However, the integration of oral health data within this digital ecosystem remains limited, particularly in terms of structured risk assessment and its translation into actionable healthcare or insurance decisions.

Risk-based approaches in healthcare, particularly those supported by data-driven models, have gained increasing importance in improving preventive care and optimizing resource allocation [5–7]. In recent years, artificial intelligence (AI) and machine learning have demonstrated significant potential in predicting disease risk and supporting clinical decision-making in healthcare [8–10]. However, the application of such approaches in oral health, especially within large-scale digital health frameworks like ABDM, remains underexplored.

The ABHA-O-SHINE (Ayushman Bharat Health Account – Oral Smart Health Insurance Neural Engine) framework is proposed to address this gap by integrating oral health risk assessment with insurance prioritization, while being designed for future AI-based automation. The present study aims to develop and pilot test this framework using a composite risk scoring system incorporating behavioral and clinical parameters, with the long-term goal of enabling scalable, AI-assisted oral health integration within the ABDM ecosystem.

## MATERIALS AND METHODS

### Study Design and Setting

A cross-sectional pilot study was conducted among patients attending the outpatient department (OPD) of Swargiya Dadasaheb Kalmegh Smruti Dental College and Hospital, Nagpur, Maharashtra, India.

### Study Population and Sample Size

A total of 126 participants were included in the study. Participants were selected randomly from the OPD population, based on predefined inclusion and exclusion criteria. The sample size was considered adequate for a pilot study aimed at feasibility assessment and preliminary validation of the ABHA-O-SHINE framework.

### Inclusion and Exclusion Criteria

Individuals aged 18 years and above who were willing to participate and provided informed consent were included in the study. Patients with systemic conditions requiring emergency care, those unable to undergo oral examination, and individuals unwilling to participate were excluded.

### Ethical Considerations

Ethical approval for the study was obtained from the Institutional Ethical Review Board of Swargiya Dadasaheb Kalmegh Smruti Dental College and Hospital, Nagpur. All participants were informed about the purpose of the study, and written informed consent was obtained prior to data collection. Confidentiality of participant data was strictly maintained throughout the study.

### Data Collection

Data collection was carried out using a structured proforma incorporating both questionnaire-based and clinical examination components. The clinical assessment was performed using the World Health Organization (WHO) Oral Health Assessment Form (2013) [11].

### The following parameters were recorded

- Demographic details (age, gender, occupation)
- Behavioral factors (tobacco usage)
- Clinical parameters including Decayed, Missing, and Filled Teeth (DMFT) index and Community Periodontal Index (CPI)

All examinations were conducted under standard clinical conditions using sterilized instruments.

### Development of ABHA-O-SHINE Risk Score

A composite oral health risk score ranging from 0 to 14 was developed by integrating behavioral and clinical parameters. Tobacco usage was coded uniformly (0 = no use, 1 = single form, 2 = multiple forms). Clinical findings from DMFT and CPI were incorporated into the scoring system based on severity.

Based on the total score, participants were categorized into:

- Low Risk: 0–4
- Moderate Risk: 5–9
- High Risk: 10–14

### Insurance Priority Categorization

Based on the risk categorization, participants were further stratified into insurance priority levels:

- Low Priority: Low-risk group
- Medium Priority: Moderate-risk group
- High Priority: High-risk group

This stratification forms the basis for integration with future insurance decision-making models within the ABHA framework.

### AI Integration Framework (Conceptual)

Although the present pilot study was conducted using manual data collection and scoring, the ABHA-O-SHINE framework is designed to be AI-compatible. The future model envisions integration of machine learning algorithms trained on structured datasets and intraoral images, enabling automated risk prediction and insurance prioritization through the ABHA-linked digital health ecosystem.

### Statistical Analysis

Data were entered into Microsoft Excel and analyzed using appropriate statistical software. Descriptive statistics were used to summarize the data in terms of frequencies, percentages, mean, and standard deviation.

### Inferential statistics included

- Chi-square test to assess association between categorical variables such as tobacco use and risk category
- Linear-by-linear association test to evaluate trend across risk categories
- Spearman’s rank correlation coefficient to assess correlation between risk score and clinical parameters (DMFT and CPI)
- Binary logistic regression analysis to identify predictors of high-risk categorization A p-value of less than 0.05 was considered statistically significant.

## RESULTS

A total of 126 participants-66 males and 60 females were included in the study. Analyses were performed on complete and valid entries for respective variables. The mean age of participants was 36.16 ± 8.42 years.

**Table 1.** Descriptive Statistics of Key Continuous Variables.

| Variable | Mean ± SD |
| --- | --- |
| Age | $36.16 \pm 8.42$ |
| DMFT | $4.28 \pm 3.53$ |
| Total Risk Score | $7.52 \pm 2.12$ |

The distribution of continuous variables demonstrated moderate variability, with risk scores showing relatively consistent clustering.

### Risk Category Distribution

The composite ABHA-O-SHINE risk score ranged from 0 to 14-The total risk score ranged from 0 to 14, comprising intervention need (0–4), tobacco use (0–2), sugar intake (0–2), DMFT score (0–3), periodontal status (0–2), and oral lesion presence (0–1), (The scoring weights were derived based on clinical relevance and existing epidemiological evidence, and were structured to ensure simplicity, interpretability, and applicability in primary care settings.).

Based on categorization: The mean ABHA O-SHINE risk score was 7.52 ± 2.12

- Low Risk (0–4): 8 participants (6.3%)
- Moderate Risk (5–9): 95 participants (75.4%)
- High Risk (10–14): 23 participants (18.3%)

**TABLE 2:**
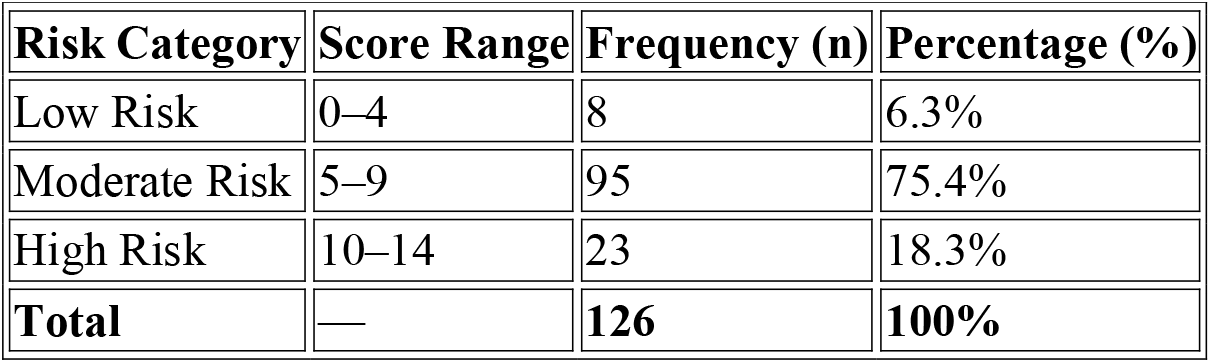
Risk Category Distribution (n = 126)

| Risk Category | Score Range | Frequency (n) | Percentage (%) |
| --- | --- | --- | --- |
| Low Risk | 0–4 | 8 | 6.3% |
| Moderate Risk | 5–9 | 95 | 75.4% |
| High Risk | 10–14 | 23 | 18.3% |
| **Total** | — | **126** | **100%** |

The majority of participants were categorized under moderate risk, followed by high risk, indicating a considerable burden of oral health risk in the study population.

### Behavioral Risk Factors

Tobacco Usage:

Tobacco consumption was reported by a substantial proportion of participants

- Non-users: 62 (49.2%)
- Single form users: 16 (12.7%)
- Multiple form users: 48 (38.1%)

**TABLE 3:** Tobacco Usage Distribution.

| Tobacco Usage | Score | Frequency (n) | Percentage (%) |
| --- | --- | --- | --- |
| Non-users | 0 | 62 | 49.2% |
| Single form users | 1 | 16 | 12.7% |

| **Tobacco Usage** | **Score** | **Frequency (n)** | **Percentage (%)** |
| --- | --- | --- | --- |
| Multiple form users | 2 | 48 | 38.1% |
| **Total** | — | **126** | **100%** |

**TABLE 4:** Association between Tobacco Use and Risk Category.

| **Tobacco Use** | **Low Risk** | **Moderate Risk** | **High Risk** | **Total** |
| --- | --- | --- | --- | --- |
| Non-users | 7 | 55 | 0 | 62 |
| Single form users | 1 | 12 | 3 | 16 |
| Multiple users | 0 | 28 | 20 | 48 |
| **Total** | **8** | **95** | **23** | **126** |

Cross-tabulation showed:

- Among non-users, the majority were in the moderate-risk group (55), with no participants in the high-risk category
- Among single-form users, 3 participants were in the high-risk category
- Among multiple-form users, 20 participants were in the high-risk category

Chi-square analysis demonstrated a highly significant association between tobacco use and risk category (χ^2^ = 34.47, p < 0.001), with a clear dose-response relationship. Notably, none of the non-users were classified under high-risk, whereas a significant proportion of multiple-form users were categorized as high risk.

Sugar Consumption: Sugar intake, assessed based on frequency in the past 24 hours, contributed to the composite risk score. A higher frequency of sugar consumption was observed predominantly among participants in moderate and high-risk categories, indicating its role as a contributory behavioral factor.

### Oral Hygiene Practices

Brushing frequency revealed that:

- 33 participants brushed once daily (26.2%)
- 93 participants claimed to brushed twice daily (73.8%)

**TABLE 5:** Brushing Frequency.

| **Brushing Frequency** | **Frequency (n)** | **Percentage (%)** |
| --- | --- | --- |
| Once daily | 33 | 26.2% |
| Twice daily | 93 | 73.8% |
| **Total** | **126** | **100%** |

Despite a majority reporting twice-daily brushing, clinical findings indicated disparities in oral hygiene status, suggesting ineffective oral hygiene practices.

### Clinical Parameters

#### Oral Hygiene Status

Based on clinical examination:

- Good oral hygiene: 24 participants (19.0%)
- Fair oral hygiene: 54 participants (42.9%)
- Poor oral hygiene: 45 participants (35.7%)

**TABLE 6:** Oral Hygiene Status.

| **Oral Hygiene Status** | **Frequency (n)** | **Percentage (%)** |
| --- | --- | --- |
| Good | 24 | 19.0% |
| Fair | 54 | 42.9% |

| Oral Hygiene Status | Frequency (n) | Percentage (%) |
| --- | --- | --- |
| Poor | 48 | 38.1% |
| **Total\*** | **126** | **100%** |

A higher prevalence of fair to poor oral hygiene was observed, correlating with increased risk scores.

#### Periodontal Status

Periodontal assessment showed that participants with periodontal pockets (>4 mm) were more likely to be categorized under higher risk groups. Spearman’s correlation revealed a strong positive correlation between periodontal status and total risk score (ρ = 0.661, p < 0.001), indicating periodontal disease as a major contributor to overall risk.

#### Dental Caries Experience (DMFT)

The mean DMFT score was 4.28 ± 3.53. Participants with higher DMFT scores were more frequently observed in moderate and high-risk categories.

A moderate positive correlation was found between DMFT and total risk score (ρ = 0.347, p < 0.01), suggesting that cumulative caries experience significantly influences risk categorization.

**TABLE 7:** Correlation Analysis.

| Parameter | Spearman's $\rho$ | p-value | Interpretation |
| --- | --- | --- | --- |
| DMFT vs Risk Score | 0.347 | <0.01 | Moderate correlation |
| Periodontal vs Risk | 0.661 | <0.001 | Strong correlation |

#### Oral Mucosal Lesions

Oral lesions associated with tobacco use were observed in 16 participants (12.7%). These participants were predominantly clustered in the high-risk category, indicating a strong association between lesion presence and elevated risk.

Healthcare Utilization Patterns

Dental Visit History

- First-time visitors: 28 (22.6%)
- Second visit: 32 (25.8%)
- Third or more visits: 66 (52.4%)

**TABLE 8:** Dental Visit History.

| Visit Frequency | Frequency (n) | Percentage (%) |
| --- | --- | --- |
| First visit | 28 | 22.6% |
| Second visit | 32 | 25.8% |
| Third or more visits | 66 | 52.4% |
| **Total** | **126** | **100%** |

A higher proportion of repeat visits suggests an existing burden of untreated or recurring oral health conditions.

Treatment Needs

Assessment of treatment requirements revealed:

- Immediate referral (advanced care): 5 (4.0%)
- Urgent treatment due to pain: 29 (23.0%)
- Scaling/minimal treatment: 69 (54.8%)
- Preventive care only: 17 (13.5%)
- No treatment required: 6 (4.7%)

**TABLE 9:** Treatment Needs Assessment.

| **Treatment Need** | **Frequency (n)** | **Percentage (%)** |
| --- | --- | --- |
| Immediate referral | 5 | 4.0% |
| Urgent treatment (pain) | 29 | 23.0% |
| Scaling/minor treatment | 69 | 54.8% |
| Preventive care only | 17 | 13.5% |
| No treatment required | 6 | 4.7% |
| **Total** | **126** | **100%** |

These findings indicate that a significant proportion of participants required active dental intervention, reinforcing the need for early risk identification.

Socioeconomic Distribution

Based on occupation:

- Unskilled/Semiskilled workers: 62 participants (49.2%)
- Skilled/Highly skilled workers: 64 participants (50.8%)

**TABLE 10:** Socioeconomic Status (Occupation)

| **Occupational Category** | **Frequency (n)** | **Percentage (%)** |
| --- | --- | --- |
| Unskilled/Semiskilled | 62 | 49.2% |
| Skilled/Highly Skilled | 64 | 50.8% |
| **Total** | **126** | **100%** |

Participants from lower occupational categories were more frequently observed in moderate and high-risk groups.

ABHA Awareness and Utilization

- Awareness of ABHA: 63 out of 126 participants (50%)
- Possession of ABHA account: 34 (26.9%)
- Active usage of ABHA at healthcare facility: 9 (7.3%)

**TABLE 11:** ABHA Awareness and Utilization.

| **Parameter** | **Frequency (n)** | **Percentage (%)** |
| --- | --- | --- |
| Unaware of ABHA | 63/126 | 50% |
| Aware of ABHA | 63/126 | 50% |
| Have ABHA account | 34/126 | 26.9% |
| Actively using ABHA | 9/126 | 7.1% |

Despite moderate awareness, actual utilization of ABHA services was found to be low.

Insurance Priority Distribution

#### Based on risk stratification

- Low Priority: 8 (6.3%)
- Medium Priority: 95 (75.4%)
- High Priority: 23 (18.3%)

**TABLE 12:** Insurance Priority Distribution.

| **Insurance Priority** | **Frequency (n)** | **Percentage (%)** |
| --- | --- | --- |
| Low Priority | 8 | 6.3% |
| Medium Priority | 95 | 75.4% |
| High Priority | 23 | 18.3% |

| Insurance Priority | Frequency (n) | Percentage (%) |
| --- | --- | --- |
| Total | 126 | 100% |

This distribution demonstrates the applicability of the ABHA-O-SHINE framework in categorizing individuals into structured insurance priority levels.

#### Logistic Regression Analysis

Binary logistic regression was explored; however, model stability was limited due to an unfavourable events-per-variable ratio and quasi-complete separation. These conditions resulted in non-convergent estimates, precluding reliable interpretation. This limitation reflects the strength of observed associations and highlights the need for larger datasets or advanced analytical approaches such as penalized regression or machine learning models.

#### Despite this, descriptive and inferential analyses consistently demonstrated that

- Tobacco use
- Higher DMFT scores
- Increased periodontal pocket depth

were strongly associated with high-risk categorization.

### Summary of Findings

The study demonstrated significant associations between behavioral factors (tobacco use, sugar intake), clinical parameters (DMFT, periodontal status, oral lesions), and composite risk score. Periodontal status emerged as the strongest clinical correlate of risk, followed by tobacco exposure and caries experience. Additionally, a high burden of treatment need and low utilization of digital health infrastructure (ABHA) were observed. The ABHA-O-SHINE framework effectively stratified individuals into clinically meaningful risk and insurance categories, supporting its feasibility for integration into the digital health ecosystem.

## DISCUSSION

Oral health continues to represent a significant yet often under-prioritized component of global health, particularly in low- and middle-income countries where the burden of preventable oral diseases remains high [1]. The present study introduces and evaluates the ABHA-O-SHINE composite index as a multidimensional tool for oral health risk stratification by integrating behavioral, clinical, and digital health parameters within a single framework.

The findings of this study demonstrate that a substantial proportion of participants were categorized under moderate to high-risk groups, reflecting a considerable underlying burden of oral disease. This is consistent with previous epidemiological evidence suggesting that oral diseases are highly prevalent and frequently untreated in community settings, particularly among populations with varied socioeconomic backgrounds [2].

A key observation in the present study was the strong association between tobacco use and increased risk categorization, with a clear dose-response relationship. Participants using multiple forms of tobacco were disproportionately represented in the high-risk category, while no high-risk individuals were observed among non-users. This finding is in alignment with well-established evidence linking tobacco exposure to periodontal destruction, oral mucosal lesions, and increased risk of potentially malignant disorders [3,4]. The clustering of oral lesions among tobacco users in the present study further strengthens this association.

Sugar consumption, incorporated as a behavioral determinant within the composite score, also contributed to elevated risk categorization. Frequent sugar intake has been consistently identified as a primary etiological factor in the development of dental caries [5], and its inclusion in the ABHA-O-SHINE framework enhances its sensitivity to modifiable risk behaviours.

An important discrepancy was observed between self-reported oral hygiene practices and clinically assessed oral hygiene status. Although a majority of participants reported brushing twice daily, a significant proportion demonstrated fair to poor oral hygiene upon clinical examination. This highlights the limitations of self-reported data and reinforces the importance of objective clinical assessment, as emphasized in standard oral health survey methodologies [6].

Among clinical parameters, periodontal status emerged as the strongest correlate of the composite risk score, demonstrating a robust positive association. This finding reflects the cumulative and multifactorial nature of periodontal disease, which is influenced by behavioral, environmental, and systemic factors [7]. Similarly, DMFT scores showed a moderate but significant correlation with overall risk, indicating that cumulative caries experience remains a critical determinant of oral health status [8].

The observed treatment needs further underscore the burden of unmet oral healthcare in the study population. A considerable proportion of participants required active intervention, including scaling and urgent care, suggesting delayed healthcare utilization and progression of disease. These findings are consistent with patterns reported in population-based studies, where curative care often supersedes preventive approaches [9].

A distinctive contribution of this study is the integration of digital health parameters through the assessment of ABHA awareness and utilization. While awareness levels were moderate, actual utilization of ABHA services remained low. This gap reflects broader implementation challenges within digital health ecosystems, including issues related to accessibility, digital literacy, and system integration, as highlighted in evaluations of national digital health initiatives [10]. By incorporating ABHA-related variables, the ABHA-O-SHINE framework extends beyond traditional indices and aligns with the evolving paradigm of digitally integrated healthcare delivery.

Importantly, the framework demonstrated effective stratification of individuals into insurance priority categories, suggesting its potential application in resource allocation and targeted public health planning. This multidimensional approach addresses a critical gap in existing indices, which are often limited to clinical parameters and fail to capture behavioral and systemic determinants of health [2,7].

Binary logistic regression was explored to identify independent predictors of high-risk categorization; however, model stability was limited. The relatively small number of high-risk cases resulted in an unfavourable events-per-variable ratio, and the presence of zero-count cells (notably the absence of high-risk individuals among tobacco non-users) led to quasi-complete separation. These conditions are known to produce unstable or non-convergent estimates in conventional regression models. From a methodological standpoint, this limitation reflects the strength of the observed associations rather than a weakness of the dataset, as the near-perfect separation of certain variables indicates a strong underlying relationship.

The ABHA O-SHINE index demonstrates strong potential for integration into digital health ecosystems. With the increasing convergence of clinical decision-making and artificial intelligence, such structured risk assessment tools can serve as foundational inputs for predictive modeling, enabling personalized and cost-effective care pathways.[12] From a forward-looking perspective, this limitation also highlights an important opportunity. The multidimensional dataset generated through the ABHA-O-SHINE framework is well-suited for advanced analytical approaches, including machine learning and artificial intelligence (AI)-based predictive modeling. Unlike traditional regression, AI models can effectively handle nonlinear relationships, complex interactions, and sparse data structures. Integration of such models could enable real-time risk prediction, dynamic risk scoring, and personalized intervention planning. Furthermore, coupling AI-driven risk stratification with digital health infrastructure such as ABHA has the potential to facilitate automated insurance prioritization, optimize resource allocation, and support decision-making at both clinical and policy levels.

It is important to note that the present study utilized manually recorded data and conventional analytical methods; however, it establishes a strong foundational dataset for future AI-driven applications. The transition from manual composite scoring to automated predictive systems represents a natural and scalable evolution of this framework.

The study is not without limitations. The cross-sectional design restricts causal inference, and self-reported behavioral variables may be subject to reporting bias. Additionally, the sample size, particularly within the high-risk category, limited the application of multivariable modeling techniques. Future studies should focus on larger, longitudinal datasets and explore the integration of AI-based analytical approaches to enhance predictive accuracy and scalability.

In conclusion, the ABHA-O-SHINE framework represents a novel and comprehensive approach to oral health risk assessment by integrating behavioral, clinical, and digital health determinants. Its ability to stratify individuals into clinically meaningful and policy-relevant categories highlights its potential for application in preventive dentistry, public health planning, and digital health integration. With further validation and technological integration, this model holds promise as a scalable tool for advancing personalized and population-level oral healthcare delivery.

To the best of our knowledge, no existing framework integrates oral health risk scoring with digital health identity and insurance prioritization within the ABHA ecosystem.

## CONCLUSION

The present study demonstrates that the ABHA-O-SHINE framework is a feasible and effective multidimensional tool for oral health risk stratification. By integrating behavioral factors, clinical parameters, and digital health indicators, the model successfully categorized individuals into clinically meaningful risk and insurance priority groups.

The findings highlight a substantial burden of moderate to high oral health risk, driven primarily by modifiable factors such as tobacco use, sugar consumption, and inadequate oral hygiene practices, alongside significant clinical contributors including periodontal disease and dental caries. The observed gap between awareness and utilization of ABHA further underscores the need for strengthening digital health implementation at the ground level.

Although conventional regression modeling was limited by data distribution characteristics, the strong associations observed across variables reinforce the robustness of the composite framework. Importantly, the structured and multidimensional nature of the dataset provides a strong foundation for future integration of artificial intelligence–based predictive models, enabling real-time risk assessment and optimized insurance prioritization.

With further validation and scalability, the ABHA O-SHINE model holds significant potential as a practical tool for preventive dentistry and public health planning, while enabling integration into national digital health ecosystems and facilitating the transition toward high-performance, AI-assisted healthcare systems.

## Data Availability

The datasets generated and/or analysed during the current study are not publicly available due to institutional policies and participant confidentiality but are available from the corresponding author on reasonable request.

## DECLARATIONS

### Ethics approval and consent to participate

Ethical approval for this study was obtained from the Institutional Review Board of Swargiya Dadasaheb Kalmegh Smruti Dental College, Nagpur, India. The study was conducted in accordance with the ethical principles outlined in the Declaration of Helsinki. Written informed consent was obtained from all participants prior to their inclusion in the study. Participation was voluntary, and participants were informed of their right to withdraw at any stage without any consequences.

### Consent for publication

Not applicable.

### Authors’ contributions

YS conceptualized the study, designed the methodology, performed data collection, conducted analysis, and drafted the manuscript.

LS contributed to data validation, interpretation of results, and critical revision of the manuscript. All authors read and approved the final manuscript.

## Acknowledgements

The authors would like to thank the participants for their cooperation and the institutional support provided by Swargiya Dadasaheb Kalmegh Smruti Dental College, Nagpur for facilitating this study.

